# Characterization of Long COVID among U.S. Medicare Beneficiaries using Claims Data

**DOI:** 10.1101/2023.03.07.23286107

**Authors:** Yun Lu, Arnstein Lindaas, Hector S Izurieta, Myrna Cozen, Mikhail Menis, Xiangyu Shi, Whitney Steele, Michael Wernecke, Yoganand Chillarige, Jeffrey A Kelman, Richard A Forshee

**Affiliations:** Center for Biologics Evaluation and Research, Food and Drug Administration, Silver Spring, MD, USA; Acumen LLC, Burlingame, CA, USA; Centers for Medicare & Medicaid Services, Washington, DC, USA

**Keywords:** long COVID, post-COVID conditions, Medicare beneficiaries, claims data, real-world data

## Abstract

This retrospective study utilized healthcare claims data to investigate the incidence, patient demographics, and concurrent diagnoses associated with long COVID in the U.S. Medicare population. Nearly 194,000 (0.6%) beneficiaries had post-COVID condition diagnoses, with higher rates among nursing home residents. Of those medically attended for COVID-19, 3-5% were diagnosed with post-COVID conditions. We observed minimal demographic differences between those with and without long COVID. When comparing diagnoses concurrent with long COVID and COVID-19, certain codes (G72 and J84) for myopathies and interstitial pulmonary diseases were disproportionately present with long COVID.

## BACKGROUND

Evidence has emerged suggesting that coronavirus disease 2019 (COVID-19) can have significant long-term sequelae [1-7]. Many individuals with a history of COVID-19 report symptoms weeks or months after their initial infection; a growing body of literature substantiates these reports and points to a broad range of symptoms and conditions that may emerge [3-6]. These persistent symptoms are commonly referred to as long COVID or post-COVID conditions [8]. While a standardized case definition of long COVID is still evolving, the broad range of sequelae experienced by patients subsequent to initial COVID-19 infection highlights the need to elucidate the natural history of this phenomenon across different populations.

This study aims to descriptively summarize the occurrence of long COVID among Medicare beneficiaries, who represent the vast majority of the older population in the U.S.

## METHODS

We utilized Medicare enrollment data and Fee-for-Service claims submitted between April 1, 2020 and May 21, 2022. For claims to be included, both the submission and service dates must fall in the date range. Medical conditions were assessed using *International Classification of Diseases, Tenth Revision, Clinical Modification (ICD-10-CM)* diagnosis codes. Medically attended COVID-19 cases had at least one claim with code U07.1 and post-COVID condition cases had either code B94.8 or U09.9 (eTable 1). Code B94.8 is a general code for sequelae of infectious diseases, used for post-COVID conditions prior to the introduction of the U09.9 code on October 1, 2021. This code was rarely used prior to the COVID-19 pandemic, which suggests that it was predominantly used to diagnose post-COVID conditions during the pandemic (eFigure 1). However, to reduce the impact of misclassification, we restricted the use of B94.8 to periods (August 1, 2020 to February 28, 2022) where ⩾70% of cases had a prior COVID-19 diagnosis. We also excluded <10 U09.9 claims with service start dates much earlier (on or prior to June 30, 2021) than the diagnosis code effective date.

We first assessed the rate of post-COVID condition diagnoses amongst all Medicare Fee-for-Service beneficiaries with continuous enrollment since April 1, 2020, separately for community-dwelling, nursing home, and end-stage renal disease (ESRD) populations. Next, we defined a long COVID case as a post-COVID condition diagnosis made ⩾28 days after an initial COVID-19 claim, consistent with the definition for long COVID used by the Centers for Disease Control and Prevention [8]. We compared the demographic characteristics and concurrent diagnoses of this long COVID population with those who survived for ⩾28 days following a COVID-19 diagnosis and did not develop post-COVID conditions. Among Medicare beneficiaries age 65 years or older, we also analyzed concurrent diagnoses using three-character *ICD-10-CM* codes to understand the broad categories of conditions associated with long COVID. We used concurrent diagnoses that appeared on the same claim without any cleaning for pre-existing conditions. Finally, we compared the prevalence of concurrent diagnoses between long COVID cases and COVID-19 cases without post-COVID conditions.

## RESULTS

From April 1, 2020, to May 21, 2022, 193,691 (0.6%) of 31,847,927 studied Medicare beneficiaries had been diagnosed with post-COVID conditions, as identified by the diagnosis codes on claims (eTable 2). This proportion was higher among nursing home residents (1.0%) than among community-dwelling beneficiaries (0.6%). Most beneficiaries diagnosed with a post-COVID condition had a previous COVID-19 claim. Specifically, 81.0% of community-dwelling, 95.3% of nursing home, and 93.2% of ESRD beneficiaries with a post-COVID condition diagnosis had a COVID-19 diagnosis on a prior claim.

Among beneficiaries with a COVID-19 claim, 142,255 (4.5%) community-dwelling, 13,384 (2.5%) nursing home, and 3,774 (4.7%) ESRD Medicare beneficiaries were subsequently diagnosed with post-COVID conditions. Around 35% of beneficiaries received their first post-COVID condition diagnosis within 28 days of their initial COVID-19 claim (eTable 3). Among beneficiaries with COVID-19 and post-COVID condition diagnoses, 105,826 (74.4%) community-dwelling and 10,380 (77.6%) nursing home residents met our long COVID definition with at least one post-COVID condition diagnosis ⩾28 days following their first COVID-19 claim. Using this case definition, the long COVID rate for community-dwelling beneficiaries (3.4%) was higher than nursing home residents (2.0%) among beneficiaries with a prior COVID-19 claim.

There were no significant sex, age, or race differences between long COVID cases and COVID-19 cases without post-COVID conditions. Among long COVID cases in the community-dwelling population, 24.7% were 80 years or older, 55.8% were female, 86.1% were white, and 18.2% were dually eligible for Medicare and Medicaid. Examining COVID-19 cases without post-COVID conditions, 22.8% were 80 years or older, 55.2% were female, 84.0% were white, and 19.1% were dually eligible. This same trend was seen in the nursing home population, with no substantial differences between long COVID and COVID-19 cases without post-COVID conditions; the exception being dual eligibility. For nursing home beneficiaries, long COVID cases had a lower proportion of dual eligibility than COVID-19 cases without post-COVID conditions (60.3% vs. 70.3%) (Table 1).

**Table 1.** Demographic Summary for COVID-19 cases with and without Long COVID – Community-Dwelling and Nursing Home Medicare Beneficiaries of All Ages.

| Demographic Characteristics | Community Dwelling |  |  |  | Nursing Home |  |  |  |
| --- | --- | --- | --- | --- | --- | --- | --- | --- |
|  | COVID-19 Only<sup>a</sup> |  | Long COVID<sup>b</sup> |  | COVID-19 Only |  | Long COVID |  |
|  | No. | % | No. | % | No. | % | No. | % |
| **Total Beneficiaries** | 2,986,027 |  | 105,826 |  | 511,836 |  | 10,380 |  |
| **Age** |  |  |  |  |  |  |  |  |
| Age below 65 | 434,163 | 14.5 | 13,901 | 13.1 | 57,344 | 11.2 | 1,116 | 10.8 |
| Age 65-69 | 625,248 | 20.9 | 20,631 | 19.5 | 50,929 | 10.0 | 1,153 | 11.1 |
| Age 70-74 | 730,601 | 24.5 | 25,712 | 24.3 | 66,284 | 13.0 | 1,455 | 14.0 |
| Age 75-79 | 514,356 | 17.2 | 19,438 | 18.4 | 73,783 | 14.4 | 1,619 | 15.6 |
| Age 80-84 | 341,572 | 11.4 | 13,198 | 12.5 | 81,373 | 15.9 | 1,756 | 16.9 |
| Age 85-89 | 203,282 | 6.8 | 8,074 | 7.6 | 83,097 | 16.2 | 1,642 | 15.8 |
| Age 90-94 | 102,387 | 3.4 | 3,749 | 3.5 | 65,478 | 12.8 | 1,125 | 10.8 |
| Age 95+ | 34,418 | 1.2 | 1,123 | 1.1 | 33,548 | 6.6 | 514 | 5.0 |
| **Sex** |  |  |  |  |  |  |  |  |
| Female | 1,648,628 | 55.2 | 59,067 | 55.8 | 323,069 | 63.1 | 6,372 | 61.4 |
| Male | 1,337,399 | 44.8 | 46,759 | 44.2 | 188,767 | 36.9 | 4,008 | 38.6 |
| **Race/Ethnicity** |  |  |  |  |  |  |  |  |
| Asian | 40,836 | 1.4 | 1,011 | 1.0 | 7,693 | 1.5 | 146 | 1.4 |
| Black | 233,343 | 7.8 | 7,540 | 7.1 | 67,609 | 13.2 | 1,240 | 11.9 |
| Hispanic | 78,022 | 2.6 | 2,352 | 2.2 | 10,153 | 2.0 | 172 | 1.7 |
| American Indian/Alaska<br>Native | 24,718 | 0.8 | 1,032 | 1.0 | 3,023 | 0.6 | 65 | 0.6 |
| White | 2,509,157 | 84.0 | 91,096 | 86.1 | 415,043 | 81.1 | 8,597 | 82.8 |
| Other | 36,631 | 1.2 | 1,116 | 1.1 | 5,181 | 1.0 | 86 | 0.8 |
| Unknown | 63,320 | 2.1 | 1,679 | 1.6 | 3,134 | 0.6 | 74 | 0.7 |
|  | COVID-19 Only<sup>a</sup> |  | Long COVID<sup>b</sup> |  | COVID-19 Only |  | Long COVID |  |
|  | No. | % | No. | % | No. | % | No. | % |
| **Medicare Dual Eligibility** |  |  |  |  |  |  |  |  |
| Dual | 571,417 | 19.1 | 19,311 | 18.2 | 359,708 | 70.3 | 6,254 | 60.3 |
| Non-Dual | 2,414,610 | 80.9 | 86,515 | 81.8 | 152,128 | 29.7 | 4,126 | 39.7 |
| **Reason for Entering Medicare** |  |  |  |  |  |  |  |  |
| Aged in without ESRD | 2,252,609 | 75.4 | 78,203 | 73.9 | 345,883 | 67.6 | 6,931 | 66.8 |
| Aged in with ESRD | 503 | 0.0 | 36 | 0.0 | 542 | 0.1 | 14 | 0.1 |
| Disabled without ESRD | 712,844 | 23.9 | 26,565 | 25.1 | 160,521 | 31.4 | 3,302 | 31.8 |
| Disabled with ESRD | 9,988 | 0.3 | 488 | 0.5 | 2,969 | 0.6 | 73 | 0.7 |
| ESRD only | 10,082 | 0.3 | 534 | 0.5 | 1,921 | 0.4 | 60 | 0.6 |
| Missing | 1 | 0.0 | - | - | - | - | - | - |

Abbreviations: ESRD, end-stage renal disease.
<sup>a</sup> COVID-19 only refers to individuals with a COVID-19 diagnosis without a subsequent post-COVID condition diagnosis.
<sup>b</sup> Long COVID refers to individuals with a post-COVID condition diagnosis $\geq 28$ days after their first COVID-19 diagnosis.

When comparing the rate ratio of diagnoses concurrent with long COVID and COVID-19, the diagnoses disproportionately present among those with long COVID included “other and unspecified myopathies”, “other interstitial pulmonary diseases”, “pressure ulcers”, “other pulmonary heart diseases”, and various encounter reason codes typically associated with procedures (Table 2 and eTable 4). Certain codes (G72 and J84) for myopathies and interstitial pulmonary diseases were disproportionally present in both community-dwelling and nursing home residents.

**Table 2.** Concurrent Three Character *ICD-10-CM* Diagnosis Summary – COVID-19 Cases with and without long COVID among Community-Dwelling and Nursing Home Beneficiaries Aged ≥65 Years.

| Diagnosis Code<sup>a</sup> | Diagnosis Code Description | COVID-19 | Long | Rate Ratio<sup>d</sup> |
| --- | --- | --- | --- | --- |
|  |  | Only<sup>b</sup> | COVID<sup>c</sup> |  |
|  |  | Claims, % | Claims, % |  |
| **Community-Dwelling Beneficiaries** |  | **n = 13,663,828** | **n = 261,417** |  |
| G72 | Other and unspecified myopathies | 0.3 | 5.3 | 16.3 |
| Z43 | Encounter for attention to artificial openings | 0.1 | 1.6 | 13.4 |
| J84 | Other interstitial pulmonary diseases | 0.5 | 7.0 | 13.1 |
| Z93 | Artificial opening status | 0.3 | 1.5 | 5.0 |
| L89 | Pressure ulcer | 0.8 | 3.8 | 4.8 |
| R43 | Disturbances of smell and taste | 0.2 | 0.9 | 4.1 |
| M25 | Other joint disorder, not elsewhere classified | 0.7 | 3.0 | 4.1 |
| R26 | Abnormalities of gait and mobility | 2.1 | 8.6 | 4.0 |
| M62 | Other disorders of muscle | 2.5 | 10.2 | 4.0 |
| G62 | Other and unspecified polyneuropathies | 0.5 | 2.0 | 4.0 |
| R13 | Aphagia and dysphagia | 1.3 | 5.1 | 3.8 |
| I27 | Other pulmonary heart diseases | 0.6 | 2.4 | 3.8 |
| Z12 | Encounter for screening for malignant neoplasms | 0.2 | 0.9 | 3.7 |
| Z86 | Personal history of certain other diseases | 3.1 | 11.5 | 3.7 |
| Z48 | Encounter for other postprocedural aftercare | 0.2 | 0.7 | 3.7 |
| **Nursing Home Beneficiaries** |  | **n = 4,378,517** | **n = 26,411** |  |
| G72 | Other and unspecified myopathies | 0.2 | 3.5 | 18.5 |
| J84 | Other interstitial pulmonary diseases | 0.4 | 2.5 | 6.9 |
| Z43 | Encounter for attention to artificial openings | 0.4 | 2.4 | 5.7 |

| Diagnosis Code<sup>a</sup> | Diagnosis Code Description | COVID-19 | Long | Rate<br>Ratio<sup>d</sup> |
| --- | --- | --- | --- | --- |
|  |  | Only<sup>b</sup> | COVID<sup>c</sup> |  |
|  |  | Claims, % | Claims, % |  |
| Z86 | Personal history of certain other diseases | 4.2 | 18.1 | 4.3 |
| Z87 | Personal history of other diseases and conditions | 2.8 | 9.4 | 3.3 |
| Z68 | Body mass index [BMI] | 1.6 | 5.1 | 3.2 |
| Z73 | Problems related to life management difficulty | 0.3 | 1.0 | 3.1 |
| I27 | Other pulmonary heart diseases | 0.7 | 2.2 | 3.1 |
| J15 | Bacterial pneumonia, not elsewhere classified | 0.7 | 2.2 | 3.0 |
| I13 | Hypertensive heart and chronic kidney disease | 1.7 | 5.0 | 3.0 |
| Z99 | Dependence on enabling machines and devices,<br>not elsewhere classified | 3.1 | 8.6 | 2.8 |
| J43 | Emphysema | 0.6 | 1.5 | 2.7 |
| I11 | Hypertensive heart disease | 2.7 | 7.1 | 2.6 |
| M48 | Other spondylopathies | 1.0 | 2.6 | 2.6 |
| Z79 | Long term (current) drug therapy | 5.7 | 14.6 | 2.6 |

Abbreviations: *ICD-10-CM, International Classification of Diseases, Tenth Revision, Clinical Modification*
<sup>a</sup> Diagnoses were summarized using the first three characters of *ICD-10-CM* diagnosis codes.
<sup>b</sup> COVID-19 only refers to claims for individuals with a COVID-19 diagnosis without a subsequent post-COVID condition diagnosis.
<sup>c</sup> Long COVID refers to claims for individuals with a post-COVID condition diagnosis $\geq 28$ days after their first COVID-19 diagnosis.
<sup>d</sup> The rate ratio was ordered in descending order among the top 100 most frequent diagnoses on Long COVID and COVID-19 only claims.

## DISCUSSION

In this large study of the Medicare population, we found that, regardless of prior COVID-19 diagnoses, almost 194,000 (0.6%) studied Medicare beneficiaries had been diagnosed with post-COVID conditions, identified by diagnoses on health care claims. The rate was higher among nursing home residents (1.0%) compared to community-dwelling beneficiaries (0.6%), which highlights the substantial burden of COVID-19 on nursing homes.

Among beneficiaries with a prior COVID-19 claim, long COVID rates were lower for nursing home residents than community-dwelling beneficiaries. This finding could be due to the combined effect of multiple factors, including a higher likelihood to detect mild/asymptomatic COVID-19 cases in nursing homes (as residents are in daily contact with health care personnel) and a lower ascertainment of post-COVID conditions among individuals with multiple existing comorbidities. Our finding that community-dwelling beneficiaries with post-COVID conditions were around four times more likely to lack a prior COVID-19 code, with approximately 19% lacking a code, supports other studies suggesting that post-COVID conditions could occur following mild cases of COVID-19 [6, 9-12].

We found lower rates of post-COVID conditions than those in prior studies of older individuals or other populations [1, 13-15]. These lower rates could be due to cases of long COVID for which health care providers did not provide a post-COVID condition code in the diagnoses in claims; this would be more likely to occur during the earlier period of the pandemic before long COVID was widely recognized and before a specific post-COVID condition diagnosis code was available. The discrepancy in long COVID occurrence across studies may also reflect the ongoing evolution in both the definition of long COVID and its recognition among health care providers.

For beneficiaries with a COVID-19 claim, we did not find major differences by age, sex or race between those with long COVID compared to those without post-COVID condition diagnoses. This suggests that on average susceptibility to long COVID may not be affected by major population demographics in this population. Given that the post-COVID condition code can represent many conditions, future studies are needed to identify and evaluate specific long COVID outcomes and related risk factors. Our finding that dual eligibility status is only associated with long COVID among nursing home residents also requires further investigation.

Among both community-dwelling and nursing-home-residing older individuals, diagnoses found to be disproportionately associated with long COVID, as compared with COVID-19 cases without post-COVID conditions, included certain codes (G72 and J84) for myopathies and interstitial pulmonary diseases. This finding is consistent with a recent study of the Medicare Advantage population that identified a substantial increase in respiratory failure (including interstitial pulmonary diseases) following COVID-19 [13]. Our findings deserve further investigation in inferential studies.

While this study provides insights into the characterization of long COVID, there are limitations. This is a descriptive study, not designed to differentiate between underlying health conditions associated with long COVID and symptoms of long COVID. The study also relies on administrative health care data.

This large study of the Medicare population captured long COVID cases using claims data, finding only minimal demographic differences between those with and without long COVID. Our finding that the overall percentage of individuals with post-COVID conditions was higher among nursing home residents highlights the substantial burden of COVID-19 on nursing homes. Our finding that certain codes for myopathies and interstitial pulmonary diseases were differentially associated with long COVID deserves investigation in inferential studies.

## Data Availability

The Medicare claims data were granted to the authors under a Data Use Agreement (DUA). Under the terms of this agreement, the authors are not allowed to share these data. Unless prohibited by law, an individual wanting access to this data could file their own DUA with the Centers for Medicare and Medicaid Services and gain approval to have access. In order to request the data, the readers may contact ResDAC (Research Data Assistance Center) at http://www.resdac.org.

## ACKNOWLEGEMENT

We thank Derick Ambarsoomzadeh, BA, Acumen LLC for contribution to writing and review of the report; Pablo Freyria Duenas, MA, Xi Li, MS, and Yue Zhang, MS, Acumen LLC for assistance with statistical programming; and Chad Buskirk, Centers for Medicare & Medicaid Services for his excellent program management.

Author’s contributions: Yun Lu, Hector S. Izurieta, Mikhail Menis, Whitney Steele, Jeffrey A. Kelman, and Richard A. Forshee contributed to the study conception and design, data interpretation, manuscript writing and review. Arnstein Lindaas, Myrna Cozen, Xiangyu Shi, Michael Wernecke, and Yoganand Chillarige contributed to study design; data collection, analysis, and interpretation; and manuscript writing and review.

## REFERENCES

1. Bull-Otterson L, Baca S, Saydah S, et al. Post–COVID Conditions Among Adult COVID-19 Survivors Aged 18–64 and ⩾65 Years — United States, March 2020–November 2021. MMWR Morb Mortal Wkly Rep 2022; 71: 713–7.

2. Carfì A, Bernabei R, Landi F. Persistent Symptoms in Patients After Acute COVID-19. JAMA 2020; 324(6): 603–5.

3. Groff D, Sun A, Ssentongo AE, et al. Short-term and Long-term Rates of Postacute Sequelae of SARS-CoV-2 Infection: A Systematic Review. JAMA Netw Open 2021; 4(10): e2128568.

4. Nalbandian A, Sehgal K, Gupta A, et al. Post-acute COVID-19 syndrome. Nature Medicine 2021; 27(4): 601–15.

5. Logue JK, Franko NM, McCulloch DJ, et al. Sequelae in Adults at 6 Months After COVID-19 Infection. JAMA Netw Open 2021; 4(2): e210830.

6. Subramanian A, Nirantharakumar K, Hughes S, et al. Symptoms and risk factors for long COVID in non-hospitalized adults. Nature Medicine 2022; 28(8): 1706–14.

7. Al-Aly Z, Xie Y, Bowe B. High-dimensional characterization of post-acute sequelae of COVID-19. Nature 2021; 594(7862): 259–64.

8. CDC. Long COVID or Post-COVID Conditions. Available at: https://www.cdc.gov/coronavirus/2019-ncov/long-term-effects/index.html. Accessed September 21, 2022.

9. Havervall S, Rosell A, Phillipson M, et al. Symptoms and Functional Impairment Assessed 8 Months After Mild COVID-19 Among Health Care Workers. JAMA 2021; 325(19): 2015–6.

10. Boscolo-Rizzo P, Guida F, Polesel J, et al. Sequelae in adults at 12 months after mild-to-moderate coronavirus disease 2019 (COVID-19). 2021; 11(12): 1685–8.

11. van Kessel SAM, Olde Hartman TC, Lucassen PLBJ, van Jaarsveld CHM. Post-acute and long-COVID-19 symptoms in patients with mild diseases: a systematic review. Family Practice 2021; 39(1): 159–67.

12. Augustin M, Schommers P, Stecher M, et al. Post-COVID syndrome in non-hospitalised patients with COVID-19: a longitudinal prospective cohort study. The Lancet Regional Health - Europe 2021; 6: 100122.

13. Cohen K, Ren S, Heath K, et al. Risk of persistent and new clinical sequelae among adults aged 65 years and older during the post-acute phase of SARS-CoV-2 infection: retrospective cohort study. BMJ (Clinical research ed) 2022; 376: e068414.

14. Ioannou GN, Baraff A, Fox A, et al. Rates and Factors Associated With Documentation of Diagnostic Codes for Long COVID in the National Veterans Affairs Health Care System. JAMA Network Open 2022; 5(7): e2224359–e.

15. CDC. Nearly One in Five American Adults Who Have Had COVID-19 Still Have “Long COVID”. Available at: https://www.cdc.gov/nchs/pressroom/nchs_press_releases/2022/20220622.htm#:~:text=Overall%2C%201%20in%2013%20adults,to%20their%20COVID%2D19%20infection. Accessed September 21, 2022.

