## Supplementary Material for "Characterization of Long COVID among U.S. Medicare Beneficiaries using Claims Data"

eTable 1. Post-COVID Condition Diagnosis Codes

| **Diagnosis Code** | **Diagnosis Code Description** |
|---|---|
| U09.9 | Post COVID-19 condition, unspecified |
| B94.8 | Sequelae of other specified infectious and parasitic diseases |

eTable 2. Summary of the Use of the Post-COVID Condition Diagnosis Code in Medicare Fee-for-Service

| **Category** | **Medicare FFS** | | **Community Dwelling** | | **Nursing Home** | | **ESRD** | |
|---|---|---|---|---|---|---|---|---|
| | **No.** | **%[a]** | **No.** | **%** | **No.** | **%** | **No.** | **%** |
| **Total Beneficiaries** | **31,847,927** | | **28,804,470** | | **2,743,507** | | **398,572** | |
| Post-COVID Condition | 193,691 | 0.6% | 163,931 | 0.6% | 26,410 | 1.0% | 4,376 | 1.1% |
| Post-COVID Condition with Prior COVID-19 diagnosis | 161,007 | 83.1% | 132,746 | 81.0% | 25,178 | 95.3% | 4,078 | 93.2% |

Abbreviations: FFS, Fee-for-Service; ESRD, End-Stage Renal Disease

[a] The percent is calculated using the number in each row as the numerator and the number in the previous row as the denominator.

eTable 3. Summary of Time Gaps Between Post-COVID Condition and COVID-19 Diagnoses

| Category | Community Dwelling | | Nursing Home | |
|---|---|---|---|---|
| | No. | % | No. | % |
| **Total Beneficiaries** | **142,255** | | **13,384** | |
| All post-COVID condition diagnoses <28 days after first COVID-19 diagnosis | 36,429 | 25.6% | 3,004 | 22.4% |
| Post-COVID condition diagnoses <28 days and ≥28 days after first COVID-19 diagnosis | 15,836 | 11.1% | 1,810 | 13.5% |
| First post-COVID condition diagnosis ≥28 days after first COVID-19 diagnosis | 89,990 | 63.3% | 8,570 | 64.0% |

eTable 4. Concurrent *ICD-10-CM* Diagnosis Summary – COVID-19 Cases with and without Long COVID among Community-Dwelling and Nursing Home Beneficiaries Aged ≥65 Year

| **Diagnosis Code** | **Diagnosis Code Description** | **COVID-19 Only**[a] | **Long COVID**[b] | |
|---|---|---|---|---|
| | | Claims, % | Claims, % | Rate Ratio[c] |
| **Community-Dwelling Beneficiaries** | | **n = 13,663,828** | **n = 261,417** | |
| R053 | Chronic cough | 0.0% | 1.9% | 39.5 |
| Z430 | Encounter for attention to tracheostomy | 0.0% | 0.9% | 36.8 |
| R5382 | Chronic fatigue, unspecified | 0.1% | 2.1% | 28.9 |
| G7289 | Other specified myopathies | 0.0% | 0.7% | 26.1 |
| Z8616 | Personal history of COVID-19 | 0.2% | 6.0% | 24.2 |
| R0609 | Other forms of dyspnea | 0.2% | 3.6% | 22.6 |
| Z431 | Encounter for attention to gastrostomy | 0.1% | 1.1% | 19.3 |
| G7281 | Critical illness myopathy | 0.2% | 4.1% | 17.1 |
| J8410 | Pulmonary fibrosis, unspecified | 0.3% | 3.9% | 15.4 |
| Z8701 | Personal history of pneumonia (recurrent) | 0.2% | 3.4% | 14.6 |
| J849 | Interstitial pulmonary disease, unspecified | 0.2% | 2.1% | 11.0 |
| J9611 | Chronic respiratory failure with hypoxia | 0.5% | 4.2% | 8.9 |
| M5450 | Low back pain, unspecified | 0.2% | 1.2% | 8.1 |
| R4189 | Other symptoms and signs involving cognitive functions and awareness | 0.1% | 1.1% | 7.6 |
| Z7409 | Other reduced mobility | 0.2% | 1.4% | 6.1 |
| **Nursing Home Beneficiaries** | | **n = 4,378,517** | **n = 26,411** | |
| Z430 | Encounter for attention to tracheostomy | 0.0% | 1.1% | 31.5 |
| G7281 | Critical illness myopathy | 0.1% | 2.2% | 19.8 |
| Z8616 | Personal history of COVID-19 | 0.6% | 10.0% | 16.7 |
| Z8701 | Personal history of pneumonia (recurrent) | 0.2% | 2.2% | 10.9 |
| J8410 | Pulmonary fibrosis, unspecified | 0.2% | 1.6% | 10.2 |
| Z8619 | Personal history of other infectious and parasitic diseases | 0.3% | 2.7% | 8.3 |
| Z20822 | Contact with and (suspected) exposure to COVID-19 | 0.5% | 3.7% | 7.2 |
| Z431 | Encounter for attention to gastrostomy | 0.3% | 1.7% | 5.9 |
| M5450 | Low back pain, unspecified | 0.2% | 1.0% | 5.8 |
| F32A | Depression, unspecified | 0.4% | 2.0% | 5.7 |
| Z9911 | Dependence on respirator [ventilator] status | 0.3% | 1.3% | 4.8 |
| J9611 | Chronic respiratory failure with hypoxia | 0.7% | 2.9% | 4.5 |
| G928 | Other toxic encephalopathy | 0.1% | 0.3% | 4.3 |
| J9622 | Acute and chronic respiratory failure with hypercapnia | 0.3% | 1.2% | 4.1 |
| Z9981 | Dependence on supplemental oxygen | 1.2% | 4.9% | 4.1 |

[a] COVID-19 only refers to claims for individuals with a COVID-19 diagnosis without a subsequent post-COVID condition diagnosis.
[b] Long COVID refers to claims for individuals with a post-COVID condition diagnosis ≥28 days after their first COVID-19 diagnosis.
[c] The rate ratio was ordered in descending order among the top 100 most frequent diagnoses on Long COVID and COVID-19 only claims.

eFigure 1. Weekly Summary of Medical Encounters with Diagnosis Code B94.8 or U09.9

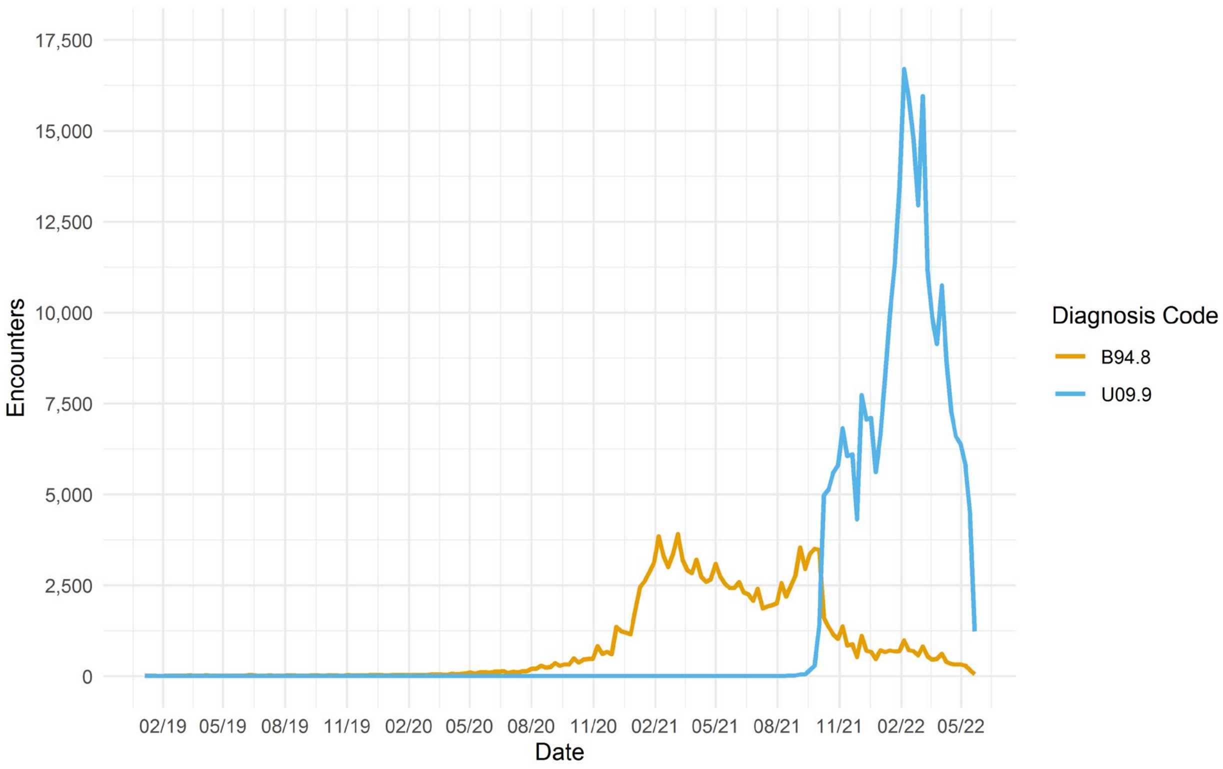
